# Hair follicle-derived epithelial sheet has potential in vitiligo treatment

**DOI:** 10.64898/2026.03.24.26349027

**Authors:** Jian Li, Jing Chen, Lichen Ling, Zheng Lin Tan, Ting Sun, Jinran Lin, Shujun Chen, Taro Uyama, Qi Zhang, Qingmei Liu, Fuyue Wu, Wenyu Wu

**Author notes:** Corresponding author. (WY.W.) (J.C.). These authors contributed equally to this work (Jian Li, Jing Chen).

## Abstract

Vitiligo is an acquired pigmentary disorder of the skin and mucus membranes. Previous study has demonstrated that autologous cultured epithelial grafts (ACEG) is an effective treatment for stable vitiligo. However, extraction of full-thickness skin might result in scar formation at donor site, which have hindered the wider application of this technology, especially for patients requiring large-area transplantation. Hair follicle as a source of keratinocyte and melanocyte, could be potential source of cells for preparation of autologous cultured sheet. Through culture system optimization, we have demonstrated maintenance of undifferentiated hair follicle-derived cells in feeder-independent culture system. After expansion, the hair follicle cells were directed to differentiate into a multi-layered, epidermis-like sheet. Cell identity, viability, purity, genomic stability, and antiseptic testing for hair follicle-derived epithelial sheet (HFES) were evaluated to ensure its safety. Immunofluorescence staining showed that basal keratinocytes were the main cell type of the autologous HFES. Optimization of culture conditions leads to increased melanocyte proliferation and functionality. Transcriptomic analysis confirmed upregulation of melanosome maturation genes. The proportions of cells are also similar to composition of cells under physiological conditions. Transplantation of HFES to depigmented areas in patients with stable vitiligo results in skin repigmentation. This technology provides a novel therapeutic option for vitiligo management.

## Introduction

Vitiligo is an acquired autoimmune disease, characterized by the selective loss of functional melanocytes, leading to well-defined white patches on the skin and mucosa. Based on clinical distribution and behavior, vitiligo is broadly classified into non-segmental vitiligo, segmental vitiligo, and unclassified vitiligo. Non-segmental vitiligo, including generalized, acrofacial, and universal subtypes, is the most prevalent and is strongly associated with autoimmune mechanisms, whereas segmental vitiligo typically presents with unilateral, dermatomal patterns and an earlier onset, suggesting distinct pathogenic features (*1, 2*). While subjective symptoms are absent for vitiligo, the course of vitiligo is long, which will result in great burden in both mental health and quality of life of the patients and their family (*3, 4*).

Currently, there are two major approaches in vitiligo treatment: (1) non-surgical approaches and (2) surgical approaches. Non-surgical treatment involves application of corticosteroids, immune modulator, e.g., Janus kinase inhibitor, vitamin D3 derivatives, and phototherapy, all aiming to modulate local immune response, thus providing the microenvironment for survival and proliferation of melanocytes. Topical corticosteroids and topical calcineurin inhibitors remain first-line therapies for localized disease, particularly effective in early or active stages (*5*). For more extensive vitiligo, phototherapy, especially narrowband ultraviolet B (NB-UVB), is considered the gold standard due to its favorable efficacy–safety profile and its ability to promote repigmentation via various mechanism, which include inhibition of proliferation of immunocytes, promotion of melanocyte migration, maturation and melanogenesis (*6, 7*). However, the efficacy rate of non-surgical methods is limited, i.e., the efficacy rate for NB-UVB phototherapy was 74.2%, while it was 29.6% for JAK inhibitor (*8*). For patients with stable refractory vitiligo, surgical approaches such as transplantation of suction blister, autologous non-cultured epidermal suspension, autologous cultured melanocytes or epithelial cell suspension have been applied with variable success. Surgical approaches have achieved an average efficacy rate of 81.01% (*9*).

However, conventional surgical interventions have the disadvantages of insufficient donor sites and difficulty in keeping cells on their places (*10*).

To address the area limitation of donor site and the problem keeping cells on the transplanted sites, we have previously developed a modified autologous cultured epithelium grafting (ACEG) to treat stable vitiligo (*11*). ACEG is tissue-engineered grafting based on cells isolated from autologous full-thickness skin from non-lesional site. The cells were cultured in a serum-free and feeder-free culture system to form an epithelial sheet without the use of exogenous matrix. ACEG were transplanted to patient’s recipient site after dermabrasion. In a clinical study conducted in 2019, we have found that the total efficacy rate of ACEG technique was 82.81% (1754/2118). The efficacy rate of ACEG is equivalent to other surgical approaches. However, with the capability of ACEG to expand approximately 100-fold of donor site, ACEG has greatly reduce the pain of patients and address the issue of limitation imposed by donor site, particularly those with large area of vitiligo lesion, when compared to conventional surgical method which can only accommodate 1:1 donor-to-recipient site area. Nevertheless, there are some disadvantages to ACEG technique. ACEG technique usually resulted in scar formation at the donor site due to full thickness skin extraction, requirement of higher-class surgical environment for donor site extraction, and occasional temporary over-pigmentation at the transplanted site. A new source to obtain the cells required for the cultivation of graft is required to further reduce the suffering of patients, and to reduce the requirement of the class of environment for surgical procedures.

Prior studies have shown that hair follicles are the sources of skin stem cells. During wounding, follicular stem cells will migrate to skin, and involved in the formation of epidermis (*12*). These stem cells reside in hair follicle bulge (*13*), which provide the potential for hair follicle regeneration and epidermis formation. Analyzing the composition of hair follicle suggested that the major components of hair follicle are keratinocytes, melanocytes, mesenchymal cells, and their stem cells, which contribute to wound healing by forming new epidermis (*14–16*). As the cellular composition of hair follicle is almost similar to epidermis, and it acts as the reservoir of skin stem cells, with appropriate culture system, hair follicles can serve as an alternative tissue source for ACEG in vitiligo treatment. This would reduce the suffering of patients and the class of environment for the extraction of tissue from donor site as follicular unit extraction results in a smaller wound (wound with diameter of < 1.0 mm) compared to full-thickness skin extraction.

Cells derived from hair follicles, particularly from outer root sheath (ORS) tissue have extensive proliferative capacity. Previous studies have shown that ORS-derived cells can be cultured into tissue-engineered skin substitute to treat various types of ulcers (*17–20*). However, these methods depend on animal-derived materials, which might result in instability and inconsistency of materials. Furthermore, melanocytes, which are important components for treatment of vitiligo lesions, were not enriched in these culture systems.

In this study, we report a hair follicle-derived epithelial sheet (HFES) cultured with a serum-free, xeno-free and feeder-free culture system. The HFES is enriched with melanocytes, which makes it a useful skin substitute to treat vitiligo lesions. The safeties of HFES will be evaluated by various testing in accordance with pharmacopeia, and exploratory clinical study was conducted to evaluate its efficacy in the treatment of vitiligo lesions.

## Results

### Establishment of a co-culture system of keratinocytes and melanocytes

In the absence of feeder cells, human epidermal stem and progenitor cells achieve effective expansion when the culture medium contains small molecules that inhibit p21-associated kinase 1/Rho-associated kinase/Myosin II and transforming growth factor β signaling under low calcium ion concentration (21).

To support growth of hair follicle-derived epidermal stem cell, a culture system containing Y-27632 (a Rho-associated kinase inhibitor) and RepSox (a transforming growth factor β receptor 1 inhibitor), insulin-like growth factor, and keratinocyte growth factor was established, yet the absence of melanocytes in this culture system is evident (Fig. 1A). This issue was addressed by supplementing 0.1 μg/mL adrenocorticotropic hormone, 0.1 μg/mL endothelin and 0.1 μg/mL stem cell factor to promote proliferation of melanocytes. This medium is known as KM + A/E/S. The presence of both melanocytes and keratinocytes was confirmed by DOPA staining with hematoxylin counterstain (Fig. 1A).

**Fig. 1.**
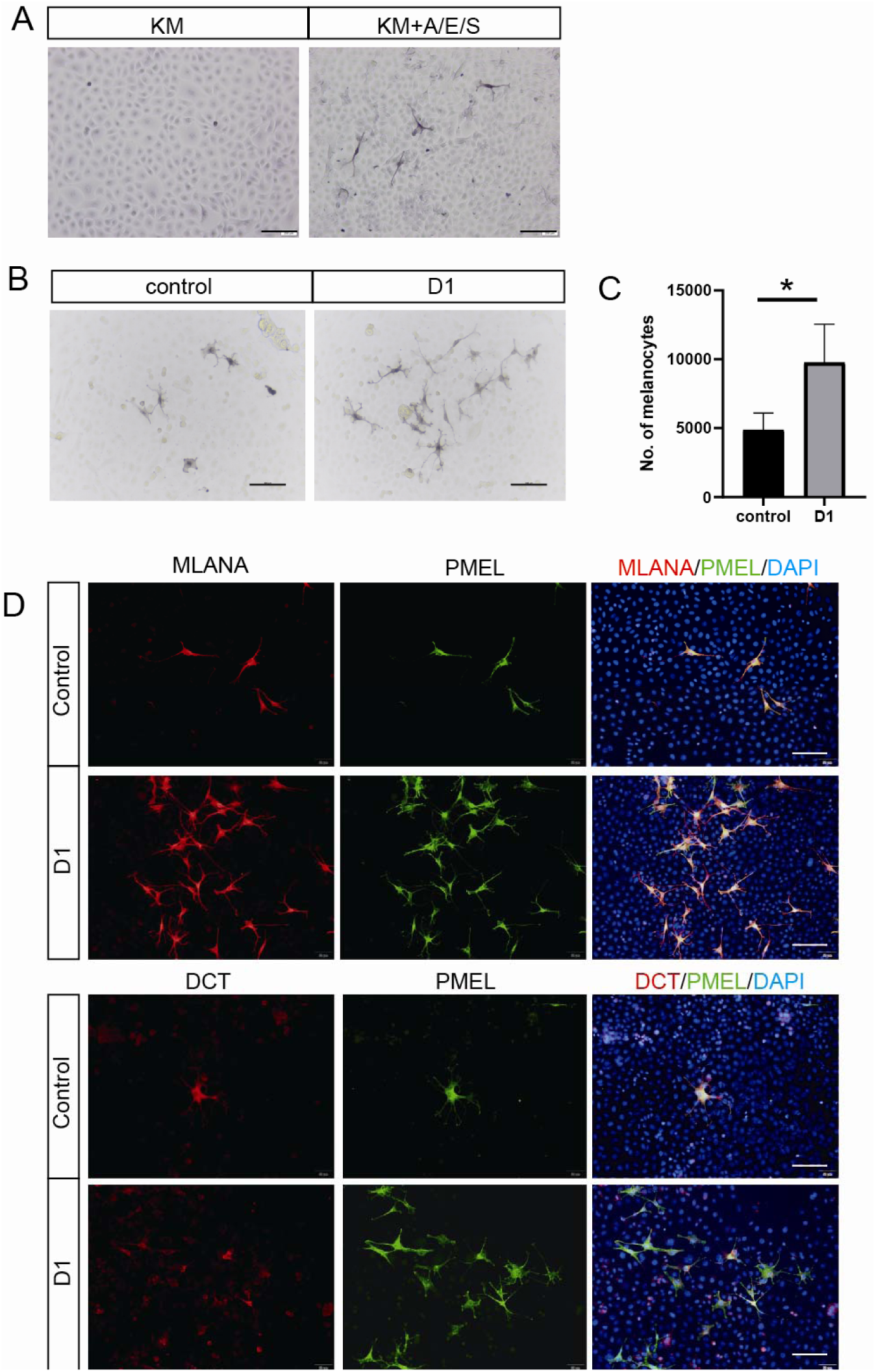
Optimization of melanocyte expansion in co-culture systems. (A) DOPA staining (brown) confirming presence of melanocytes and keratinocytes in KM and KM+A/E/S co-culture medium. Scale bar: 100 μm. B. DOPA staining of combined melanocyte stimulating factors. Scale bar: 100um C. Number of DOPA+ melanocytes per dish. (n=3, *p□<□0.05. **p□<□0.01. ***p□<□0.001. ****p□<□0.0001.). D. Immunostaining of melanocyte markers was performed on hair follicle-derived cells (n=3) under different conditions. scale bar: 100 μm.

### Optimization of the culture conditions for melanocyte proliferation

The content of melanocytes is crucial in the treatment of vitiligo lesion based on our previous study (11), therefore, it is important to ensure the optimal content of melanocytes in HFES.

Based on KM + A/E/S, we have measured the content of melanocytes and found that the number of melanocytes was < 5,000 cells per sheet, which was too low for treatment of vitiligo lesion. Therefore, it is important to optimize our culture system to improve the conditions of melanocyte proliferation.

To further increase the number of melanocytes in our culture, we investigated the influence of combination of melanocyte growth supporting factors, i.e., basic fibroblast growth factor (bFGF), α-melanocyte stimulating hormone (α-MSH), endothelin and forskolin on the content of melanocytes in the co-culture system of keratinocytes and melanocytes. As shown in Fig. S1A – S1B, we found that 0.05 μg/mL bFGF, 0.2 μg/mL α-MSH, 0.2 μg/mL endothelin and 10 μM forskolin can promote the growth of melanocytes and increase the number of melanocytes. In combination, bFGF and forskolin (B + F) group demonstrated the strongest ability to increase the number of melanocytes (Fig. S1C, D). We named this culture medium containing KM + A/E/S + B + F as “D1”. Compared to KM + A/E/S (Control), D1 significantly increased the number of DOPA^+^ melanocytes (Fig.1B, C). These proliferating melanocytes express Melan-A (MLANA), pre-melanosome protein (PMEL) and dopachrome tautomerase (DCT), suggesting that they are cells from melanocytic lineage (Fig.1D).

### Transcriptomic analysis of optimized culture medium

Plot of principal component analysis showed high dispersion of data in principal component 1 (Fig. S2A), suggesting high biological variability of sample. Yet each sample cultured in D1 was closed to their respective control, suggesting that the culture system was stable despite of the high variability of biological sample. (Fig. 2A). Among the differentially expressed genes of sample cultured with D1 compared to control, 192 genes were upregulated, including CA9, STC1, MMP9, IL11, HGF, FGF19, GPR4, and 203 genes were downregulated, including DKK1, FN1, CLDN11, HSPA8, AQP1, WNT5B, ACAN (Fig. 2B).Heatmap with group average value showed upregulation in TYRP1, PMEL, RAB38, ERBB3, RAB27A in D1 group compared to control, which suggested upregulation in melanin synthesis (Fig. 2C). No significant differences in the genes related to keratinocytes stemness and function were observed (Fig. 2D), suggesting that the function of keratinocytes did not reduce in D1. Validation with quantitative polymerase chain reaction confirmed the upregulation of TYRP1, PMEL, RAB38, RAB27A and transcription factor AP-2 alpha (TFAP2A) (Fig. 2E). These genes are involved in regulation of melanocyte differentiation, melanosome biogenesis, function, and transport (*22*). The upregulation of these genes in cells cultured with D1 medium indicates that D1 can promote maturation of melanocytes and enhance melanin production without affecting the function of keratinocytes.

**Fig. 2.**
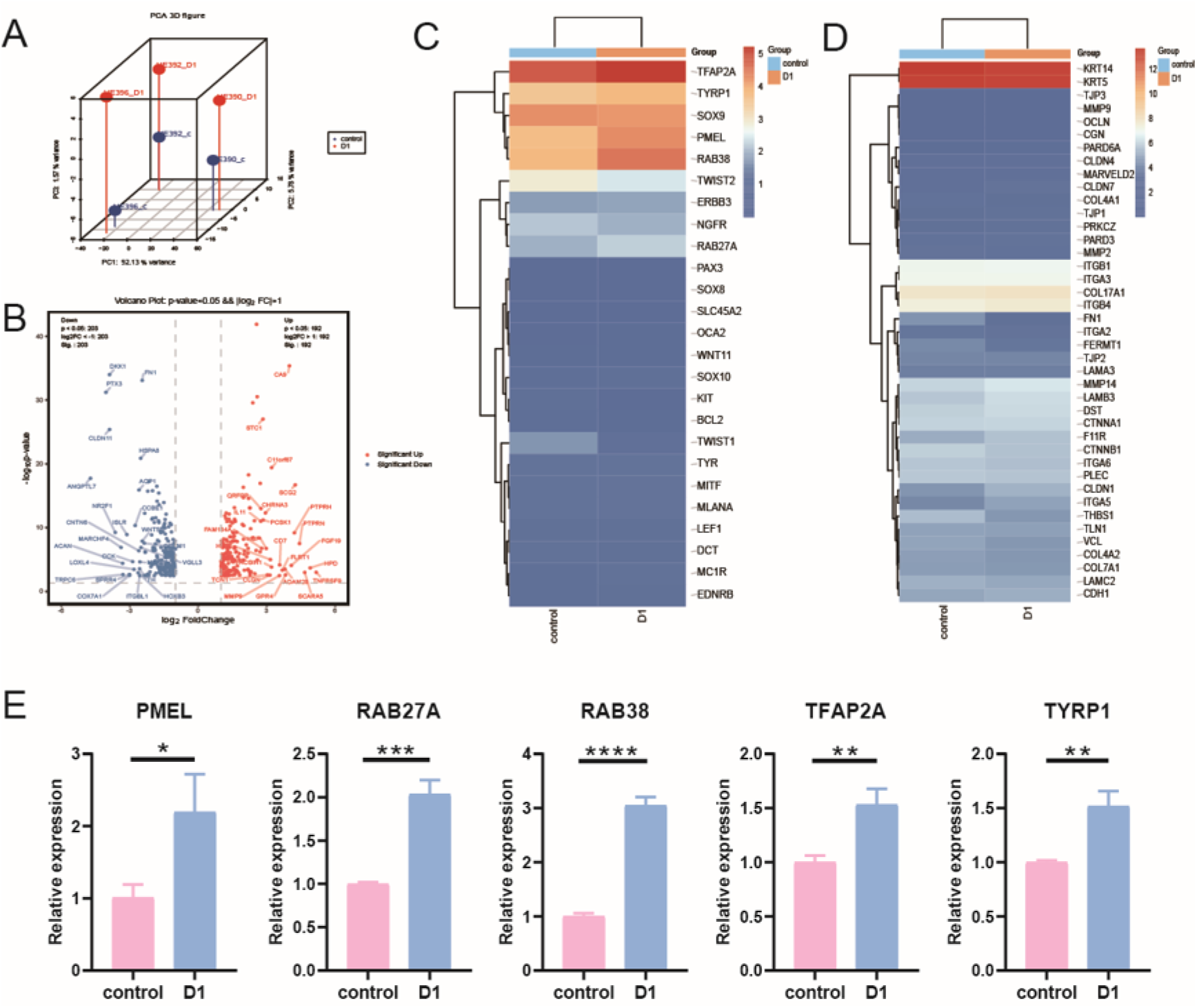
Transcriptomic analysis of optimized culture medium. (A) 3D principal-component analysis (PCA) comparing the transcriptome of control and D1. (B) Volcano Plot of differential expression gene, (C) Heatmap plotting expression of melanocyte related genes. (D) Heatmap plotting expression of epithelial cells related genes. (E) qPCR verification of melanocyte characteristic genes that show differential expression in bulk-RNA-Seq analysis (n=3, *p□<□0.05. **p□<□0.01. ***p□<□0.001. ****p□<□0.0001.)

### The hair follicle cells were expanded in vitro to obtain functional melanocytes and proliferative keratinocytes similar to the basal layer of the epidermis

After passaging, the medium was replaced with E1 medium, which is a medium significantly prolonged the in vitro survival and proliferative capacity of human epidermal keratinocytes to support the growth of hair follicle keratinocytes to 100% confluence. After expansion in E1, the cells were stained and identified by immunofluorescence staining (Fig. 3).

**Fig. 3.**
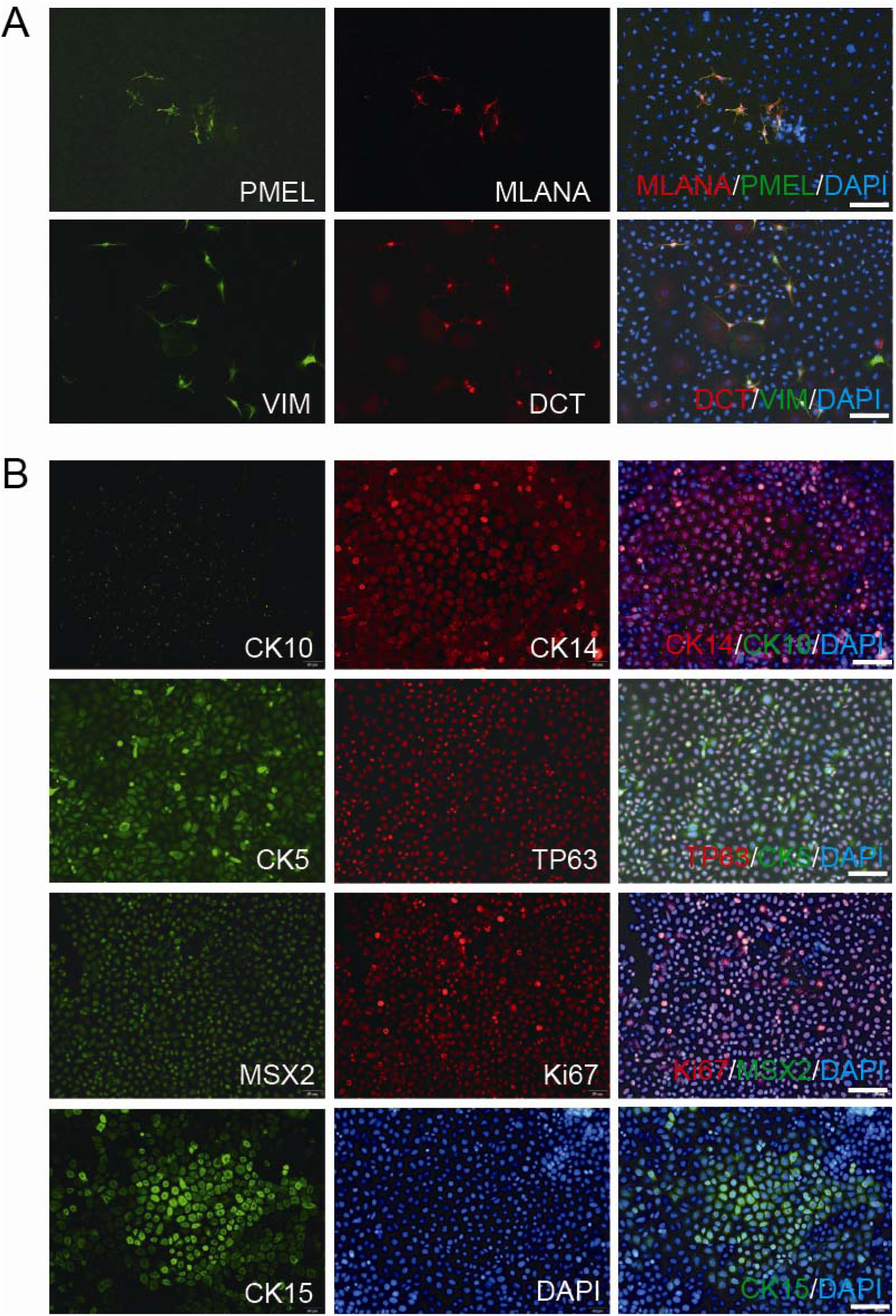
The constituent cells of hair follicle derived epithelial sheets. (A) Melanocytes markers PMEL, MelanA, DCT and Vimentin were tested on P1 cells. (B) keratinocytes markers CK10, CK14, CK5, TP63, CK15 and MSX2, Ki67 were tested on P1 cells. Scale bar: 100 μm

The melanocytes were primarily PMEL^+^ MLANA^+^ melanocytes. DCT was expressed in some of the melanocytes, exhibiting different morphology compared to those PMEL^+^ MLANA^+^ melanocytes population. Vimentin (VIM)^+^ subpopulation was also detected in our culture (Fig. 3A). DCT^+^ melanocytes were spindle-shaped, while PMEL^+^ MLANA^+^ melanocyte exhibit polydendrite morphology.

In order to deduce the tissue origin of the cultivated keratinocytes, we examined whether these cells exhibited properties of hair follicle-derived keratinocytes (Fig. 3B). All cells cultured with E1 were CK14^+^ with subpopulations of CK5^+^ and CK15^+^ cells. As these cells were proliferative in our culture medium, they are also tumor protein p63 (TP63)^+^ and Ki67^+^.

### The hair follicle-derived cells form an epidermis-like stratification epithelial sheet rich in melanocytes

High concentration of calcium ion in culture environment leads to the formation of tight junctions and desmosome, promoting withdrawal from proliferation and the assembly of organized, multilayered epidermis-like structures in vitro (*23*). In culture medium with high calcium concentration (designated as F1), keratinocytes progressively differentiated and formed stratified multilayered-structure (Fig. 4A). After cultivation in F1, cohesive multilayer sheet was detached by collagenase digestion (Fig. 4B). The detached HFES exhibited robust structural integrity, permitting repeated rinsing with PBS to eliminate residual culture medium and enzymes (Fig. 4B). Histological analysis via cryosectioning stained with hematoxylin and eosin staining revealed a stratified epidermal-like architecture in the graft, with thickness of 28.4 μm (Fig. 4C). Immunofluorescence staining of tissue sections demonstrated that cells were CK14^+^ cells, with TP63^+^ cells localized predominantly at the basal layer of HFES (Fig. 4D). The karyotypes of hair follicle-derived cells were maintained through 5th passage in D1, indicating genomic stability during serial passaging (Fig. 4E).

**Fig. 4.**
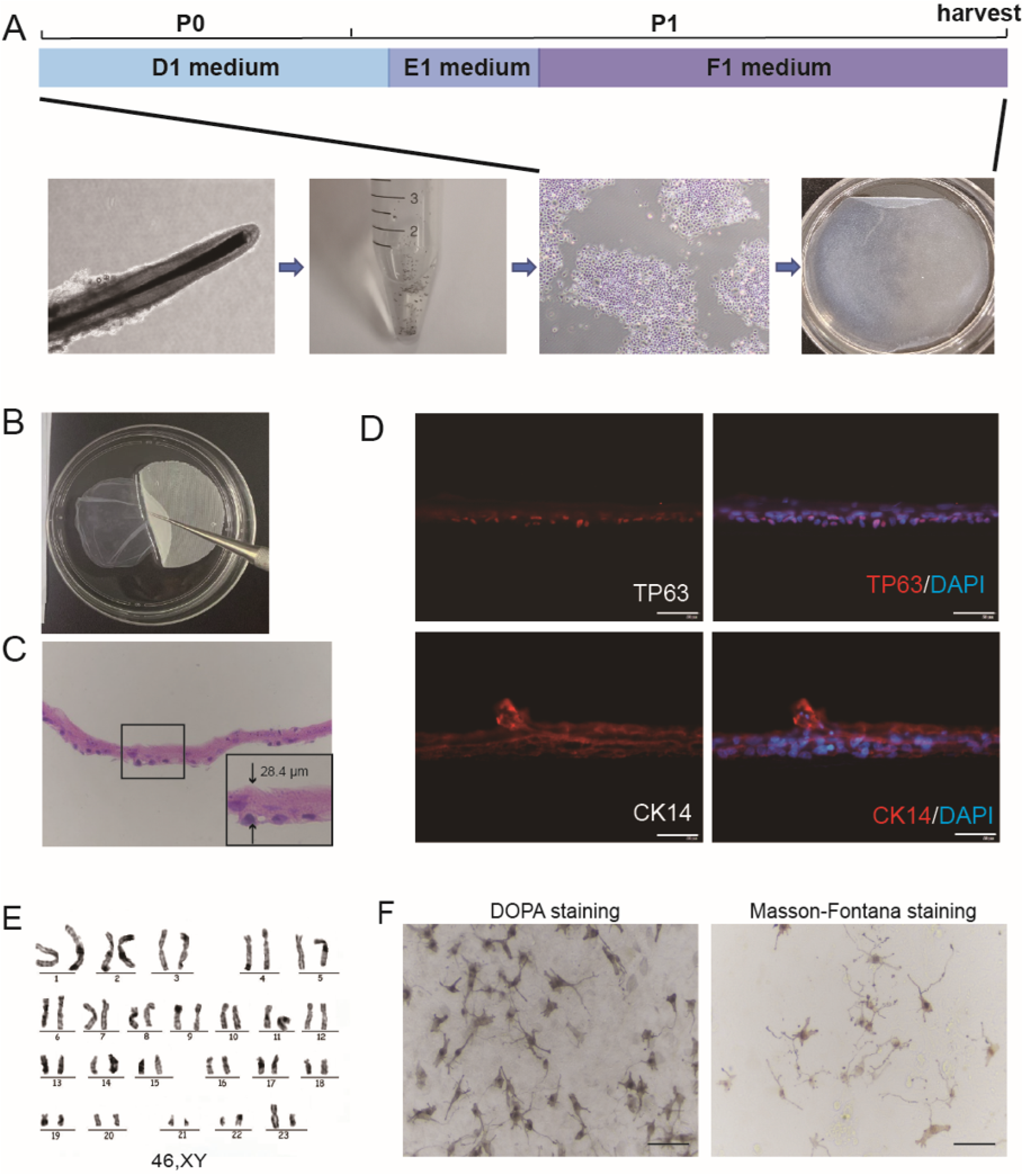
Production and evaluation of HFES. (A) Schematic showing the procedure of generation of HFES. (B) Cleaning of harvested epithelial sheet. (C) H&E staining of harvested epithelial sheet. (D) Immunofluorescence detection of epidermal basal layer markers TP63 and CK14 on the harvested epithelial grafts. scale bar: 50μm. (E) Karyotype of 5^th^ passage hair follicle cells. (F) The melanocytes in the harvested epithelial grafts were evaluated through DOPA staining and Masson-Fontana staining. scale bar: 100μm.

To evaluate the function of melanocyte in HFES, we have conducted DOPA staining and Masson-Fontana staining of HFES. DOPA staining indicates tyrosinase activity in melanocytes, indicating melanin synthetic capability; while Masson-Fontana staining indicates the density of melanin pigment in melanocytes (Fig. 4F). Both DOPA and Masson-Fontana positive melanocytes were present in HFES. Melanocyte density exhibited significant inter-sample variation, ranging from 6628 to 36978 cells per dish among specimens, contributing to an average K:M at 71.4 :1 (Table 1.). These characteristics suggest that HFES have potential in vitiligo treatment.

**Table 1.** Number of melanocytes and keratinocytes to melanocytes ratio (K:M) of HFES.

| Sample ID | No. of melanocytes per<br>HFES | K:M |
| --- | --- | --- |
| HF250 | 11680 | 73:1 |
| HF251 | 16270 | 56:1 |
| HF252 | 20538 | 49:1 |
| HF253 | 36978 | 27:1 |
| HF254 | 6628 | 152:1 |

### Single-cell transcriptomic profiling reveals dynamic compositional and functional remodeling during tissue reconstruction

After data filtering, 15,036 cells were identified from 6 samples, with 22 transcriptionally distinct populations across intact hair follicles (HFs), primary cultures (P0 cells), and epithelial sheets (sheets) (Fig. 5A). Major cell types included keratinocyte subtypes (basal keratinocytes, suprabasal keratinocytes, spinous keratinocytes, ORS keratinocytes, ORS-like keratinocytes), melanocytes, dermal papilla (DP) cells, and stromal components (immunocytes, endothelial cells, smooth muscle cells) (Fig. 5B, C). Comparative cellular distribution analysis demonstrated significant inter-sample heterogeneity: HFs exhibited niche specialization with elevated DP cells (3.53% - 6.93%) and melanocyte (1.28% - 1.43%), while HFES retained epidermal progenitors including basal keratinocytes 1 (4.32% - 11.63%) and ORS-like keratinocytes (2.88% - 5.51%). DP cells were not detected. Melanocytes formed 3.69% - 8.87% of HFES, which is higher than HFs and the result obtained from microscopy examination (Table 1.), which is 0.46% - 3.65%. P0 cells were dominated by proliferative states such as proliferating keratinocytes (77.14% - 82.38%) and cycling keratinocytes (0.24% - 0.37%), indicating adaptation to in vitro expansion (Fig. 5D).

**Fig. 5.**
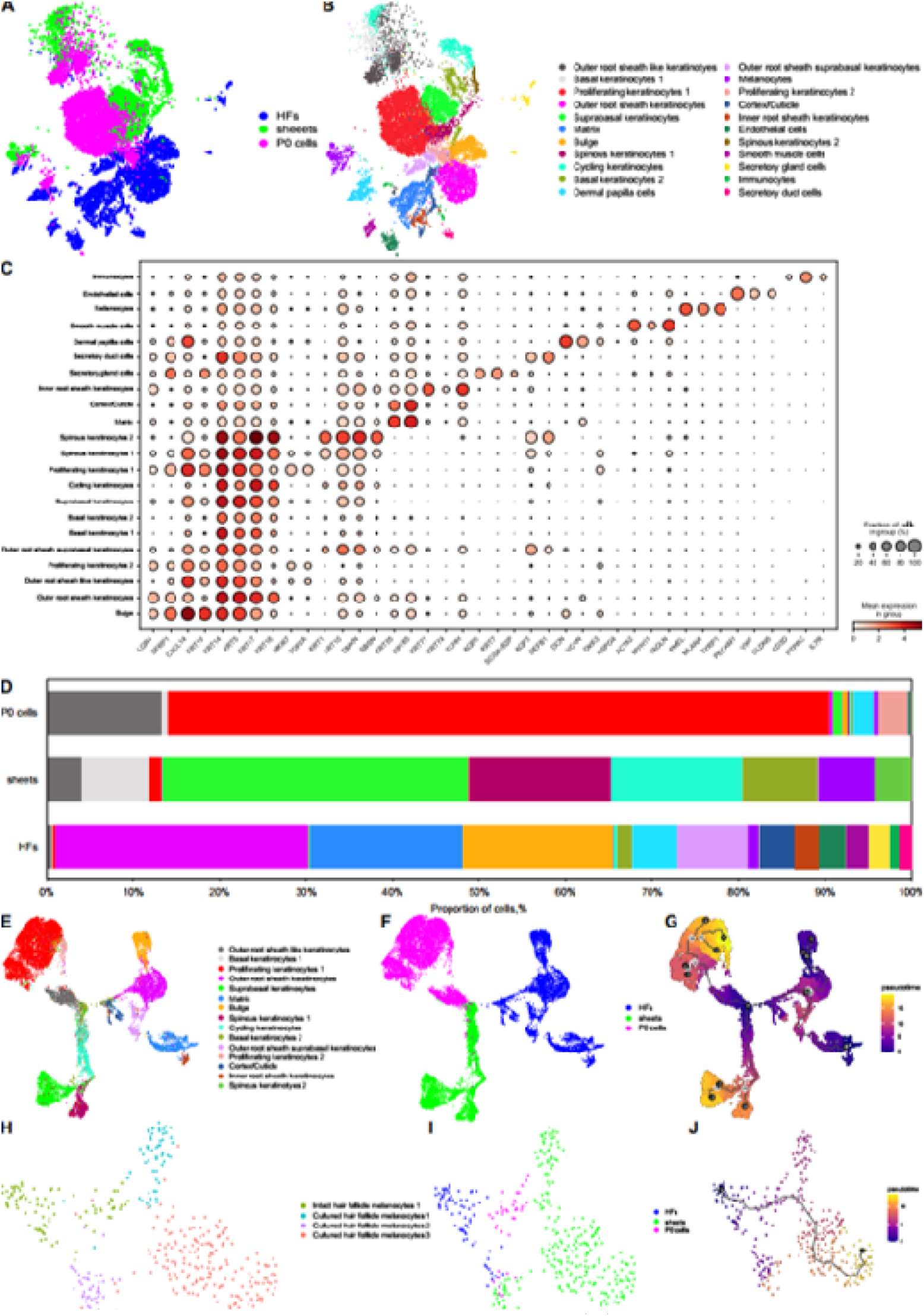
scRNA-seq analyses of HFs, sheets and P0 cells. (A) UMAP plot of data grouped by origin of samples and (B). by cell type. Each point is a cell, and different cell types are differentiated by different colors. (C) Dot plot of each cell type representing genes expressed by each cluster. (D) Proportion of cells in each cluster. UMAP plot of keratinocytes as a subcluster of A grouped by (E) cell type and (F) origin of samples. (G) Pseudotime trajectory map of keratinocytes. UMAP plot of melanocytes as a subcluster of A grouped by (H) cell type and (I) origin of samples. (J) Pseudotime trajectory map of melanocytes.

Pseudotemporal trajectory reconstruction positioned cells with higher degree of stemness as origin, which are bulge cells and hair matrix cells, with subsequent bifurcation into terminal differentiation pathways: one branch progressed toward P0 cells, while the other advanced toward HFES. HFES exhibited accelerated epidermal commitment with highest degree of differentiation, while HFs remained relatively undifferentiated (Fig. 5E-G)

Subclustering of 359 melanocytes revealed state-specific transitions. Melanocytes were clustered into 4 clusters based on their origin (Fig. 5H, I). Quantitative analysis confirmed progressive transition of melanocyte from HFs (1.28% - 1.43%) to P0 cells (0.31% - 1.00%) and proliferated into sheets (3.69% - 8.87%) (Fig. 5H-J). These findings collectively demonstrate that HFES preserve critical epidermal progenitor compartments and functionally activated melanocytes while undergoing accelerated stratification, supporting their potential as biologically competent constructs for vitiligo treatment.

### Potential of HFES in treatment of vitiligo lesions

To evaluate the real therapeutic potential of HFES for stable vitiligo, we enrolled five patients with stable vitiligo for HFES transplantation and assessed the safety and therapeutic outcomes (Table S2.). Following HFES transplantation, none of the five patients experienced infection or other adverse events. Assessment of therapeutic efficacy at transplant sites using inverse VASI scoring demonstrated the efficacy of HFES in treatment of vitiligo lesion (Fig. 6). Average repigmentation rate was 98% (Table 2.).

**Fig. 6.**
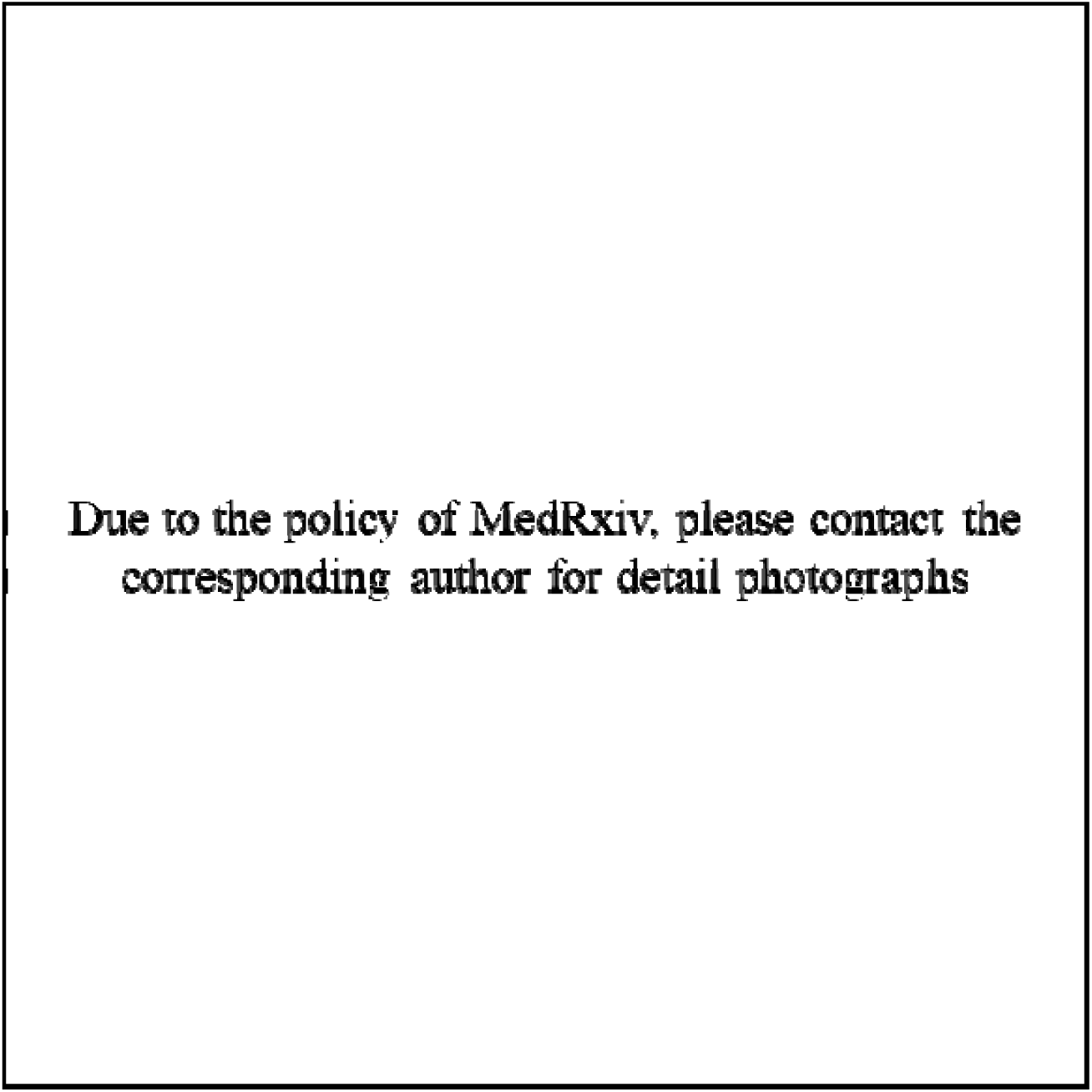
Comparison of clinical repigmentation in stable vitiligo patients after HFES treatment. (A) Case1. (B) Case2. (C) Case3. (D) Case4. (E) Case5. Grafting sites were highlight with blue lines.

**Table 2.** Inverse VASI score of HFES.

| case | HFES |
| --- | --- |
| 1 | 100% |
| 2 | 100% |
| 3 | 100% |
| 4 | 100% |
| 5 | 90% |
| mean | 98% |

## Discussion

In this study, we developed a therapeutic strategy for stable vitiligo using autologous hair follicle-derived epidermal sheets (HFES) generated under a fully defined, serum-free, xeno-free, and feeder-independent culture system. Previous work has demonstrated that hair follicle-derived keratinocytes can be propagated in medium containing 0.05–0.1 mM calcium and supplemented with epidermal growth factor, bovine pituitary extract, fetal calf serum, insulin, and cholera toxin (*24*); however, the proliferative capacity of these cultures remains limited, requiring approximately two weeks for expansion. For clinical translation, both safety and batch-to-batch consistency demand the elimination of cholera toxin and all animal-derived components, including serum and bovine pituitary extract, in favor of a chemically defined formulation. Our culture system meets these criteria through the use of regulatory-compliant, chemically defined reagents, while deliberately excluding tumorigenic compounds such as 12-O-tetradecanoylphorbol-13-acetate and 3-isobutyl-1-methylxanthine (*25–30*). By systematically comparing the effects of bFGF, α-MSH, endothelin, and forskolin on melanocytes in a co-culture setting, we established KM + A/E/S medium, which simultaneously supports the expansion of both keratinocytes and melanocytes. Further optimization enhanced melanocyte proliferation and drove a functionally mature phenotype, as evidenced by the upregulation of melanosomal maturation genes, including TYRP1, PMEL, and RAB27A. PMEL and MLANA are canonical melanocyte differentiation antigens integral to melanosome biogenesis; PMEL provides the structural fibrils of premelanosomes, while MLANA contributes to melanosome stability and melanin synthesis. DCT functions as a melanogenic enzyme whose expression correlates with pigment production in functional melanocytes (*31–33*). Notably, melanocytes maintained in D1 medium displayed a PMEL^hi^ MLANA^hi^ DCT^lo^ profile, indicative of a transit-amplifying state in which low enzymatic activity presumably prevents the premature accumulation of reactive oxygen species. This interpretation is reinforced by widespread Ki67 and TP63 expression across the population and by attenuated DOPA staining in D1-cultured melanocytes. Upon maturation into HFES, melanocytes upregulated DCT to support melanin biosynthesis. Within HFES, we identified a subset of DCT VIM human melanocytes with spindle-shaped morphology—distinct from the polydendritic appearance of mature melanocytes. Given that mesenchymal cells were not detected in the scRNA-seq data, these cells are of melanocytic lineage; their morphology and VIM expression, a marker often associated with mesenchymal stem cells, suggest that they represent melanocyte progenitor cells or migratory melanoblasts.

Keratinocytes within the hair follicle ORS constitutively express CK5, CK14, CK15, and CK17. In HFES, we detected CK14□, CK5□, and CK15□ populations, while CK10 was uniformly absent, confirming a basal keratinocyte identity. This basal/progenitor state was further corroborated by universal MSX2 expression, a transcription factor that maintains the undifferentiated status of keratinocytes. Moreover, the cultured keratinocytes were highly proliferative, as reflected by TP63 and Ki67 co-expression (*34–37*). Taken together, hair follicle-derived keratinocytes expanded efficiently while preserving basal and progenitor characteristics, whereas melanocytes concurrently progressed toward functional maturation, acquiring robust melanin-producing capacity. The resultant HFES are stratified epithelial sheets in which proliferative cells are confined to the basal layer, and CK14 is expressed throughout.

When cultured in D1 medium, the cells retained genomic stability over multiple passages, supporting the clinical safety of HFES generated in this system. DOPA staining and Masson–Fontana staining confirmed melanocyte functionality, and single-cell transcriptomic quantification revealed a melanocyte-to-keratinocyte ratio approaching that of native epidermis, underscoring the potential efficacy of HFES in vitiligo treatment.

Single-cell transcriptomics offered unprecedented resolution of the cellular dynamics underlying graft development. Dimensionality reduction and clustering of 15,036 cells revealed that HFES harbored a markedly elevated proportion of epidermal progenitors, significantly exceeding the 17.7% observed in intact hair follicles. Pseudotemporal trajectory analysis demonstrated accelerated epidermal commitment within the sheets, a process driven by calcium-mediated differentiation. The percentage of melanocytes increased from hair follicles to sheets, indicating successful melanocyte expansion. Given that an individual melanocyte is naturally surrounded by 25–40 keratinocytes to form an epidermal melanin unit, the melanocyte proportion in our sheets closely mirrors the physiological state. This finding demonstrates that our culture protocol selectively enforces a melanogenically competent phenotype essential for effective repigmentation.

An exploratory clinical study enrolled five patients with stable vitiligo, encompassing both segmental and non-segmental subtypes. The repigmentation efficacy, assessed by inverse VASI scoring, reached 98% in the treated areas. This outcome compares favorably with autologous cultured epidermal grafts (ACEG), while offering a shorter culture period (17 days for HFES vs. 20 days for ACEG) and a less invasive donor-site harvesting procedure, positioning HFES as a promising therapeutic avenue for stable vitiligo.

This study has several limitations that warrant future investigation. A randomized comparative clinical trial is needed to evaluate the efficacy of HFES against established modalities such as ACEG or narrow-band ultraviolet B phototherapy. Mechanistic analyses within such a trial should elucidate which HFES subpopulations predominate in driving repigmentation and how these cells contribute to immune modulation within vitiligo lesions, thereby achieving durable repigmentation.

In conclusion, HFES represent a therapeutically robust and minimally invasive solution for stable vitiligo. The cellular and molecular mechanisms governing repigmentation in patients will be dissected in subsequent investigations.

## Materials and Methods

### Cell Isolation and Cultivation

Follicles were harvested via motorized FUE: punch penetration, suction extraction, atraumatic retrieval with curved forceps, preserving structural integrity. Isolated hair follicles were immersed in culture medium containing 2.4 U/ml neutral protease and incubated overnight at 37°C. The tissues were then minced and digested with TrypLE™ Express (Gibco, 12604-021) for 20 – 40 minutes. The resulting suspension containing cells and tissue fragments was filtered through a 70μm cell strainer to obtain a single-cell suspension.

Trypsinized cells were collected and cultured in type IV collagen-coated dishes. Cultures were maintained in defined keratinocytes-serum free medium (DK-SFM, Gibco, 10744-019) supplemented with RepSox (Selleck, S7223), Y-27632 (Selleck, S1049), adrenocorticotropic hormone (ChinaPeptides, 12279-41-3), endothelin (ChinaPeptides, 143113-45-5), stem cell factor (Peprotech, 300-07), bFGF (Oryzogen, HYC005M01), and forskolin (Selleck, S2449) at 37°C with 5% CO2. Medium was changed every other day. Upon reaching 80% confluence, cells were passaged and cultured in E1, an Epilife medium with 60 μM calcium (Gibco, MEPI500CA)-based medium for expansion at 37°C with 5% CO2. Upon reaching 100% confluency after expansion in E1 medium, the medium was replaced with F1, a DMEM/F12-based medium (3:1 ratio) supplemented with 1.4 mM calcium, 1% non-essential amino acids (ThermoFisher Scientific, 11140050), 10 ng/mL epidermal growth factor (Oryzogen, HYC020M01), and 100 ng/mL insulin-like growth factor (Oryzogen, HYC007M01). Cells were subsequently maintained in F1 medium with daily medium changes until forming stratified epidermal sheets.

### DOPA Staining for Melanocyte Detection

Samples were fixed at room temperature for 30 min, washed with D-PBS (Basal Media, B210KJ) for 3 times, and air-dried. Fresh DOPA solution was prepared by dissolving 0.1 g DOPA (Sigma, D9628-5G) in 100ml DPBS. Samples were immersed in DOPA solution and incubated at 37°C for 6 hours protected from light. After gentle washing with distilled water, samples were completely air-dried for microscopic melanocyte evaluation.

### Masson-Fontana Staining

Cells were fixed with 4% paraformaldehyde in PBS, pH 7.4, for 20 minutes at room temperature and washed with D-PBS. Masson-Fontana Staining using a Masson-Fontana Melanin Stain Kit (Solarbio, G2032), according to the manufacturer’s instructions.

### Bulk-RNA-seq Analysis

Total RNA was extracted and treated with DNase I to remove genomic DNA. mRNA was enriched via oligo(dT) magnetic bead selection. Purified mRNA was fragmented using divalent cations under elevated temperature, followed by first-strand cDNA synthesis with random hexamer primers. Double-stranded cDNA (dsDNA) was generated through second-strand synthesis and purified using AMPure XP beads. The dsDNA underwent end repair, 3’-adenylation, and Illumina adapter ligation. Size selection (300–500 bp) was performed via bead-based purification, and final libraries were amplified by 12-cycle PCR. After quality control (Agilent 2100 Bioanalyzer, DV200 > 80%), libraries were sequenced on Illumina platforms to generate 150-bp paired-end reads.

### scRNA-seq Analysis

Tissue and cells samples were processed using scRNA sequencing platform from 10x Genomics. Single-cell suspensions were encapsulated into 35 μm microdroplets with barcoded gel beads in a microfluidic chip, followed by reverse transcription using an enhanced RT enzyme. cDNA underwent amplification (12 cycles), SPRIselect bead purification (0.6× ratio), and fragmentation. Dual-indexed libraries were constructed via end repair, adapter ligation, and index PCR (14 cycles). Final libraries were quantified by Qubit and validated for fragment distribution (350–450 bp) via Agilent 2100 Bioanalyzer. All steps used nuclease-free reagents and magnetic separation (DynaMag™). FASTQ files were processed in Cell Ranger v9.0.1 and mapped to GRCh38-2024-A transcriptome. Then, the pre-filtered gene expression matrices were imported into R Seurat package v5.3.0 for downstream analysis. Cells containing <200 genes and genes expressed in <3 cells of the data were removed, and low-quality cells with >20% of UMIs derived from the mitochondria were also excluded from further analyses. Data was normalized, scaled, the dimension was reduced, and the data was integrated. Pseudotime trajectory analysis of subclustered data was conducted with Monocle3 package (v1.4.26).

### Histological and immunohistochemical staining

Cells were seeded on collagen IV pre-coated coverslips, and the cells were cultured until cell confluence exceeded 80%. Cells on coverslips were fixed with 4% paraformaldehyde in PBS, pH 7.4, for 20 min at room temperature and washed with PBS.

HFES samples were washed with PBS and fixed with 4% paraformaldehyde in PBS, pH 7.4, for 20 min at room temperature and washed with PBS. After sucrose gradient dehydration, samples were embedded in O.C.T. Compound (Sakura Tek, 4583), 5 mm-thick sections were cut and stained in Meyer’s hematoxylin and eosin Y solutions.

For immunostaining, samples were first blocked in PBS containing 5% BSA and 0.2% Triton X-100 for 1 hour at room temperature and subsequently incubated overnight with primary antibodies at 4°C. After incubation, the solution was decanted, and the cells were washed 3 times in PBS. The secondary antibody was diluted in 1% BSA, added to the coverslip, and incubated for 1 hour at room temperature in the dark. Finally, the cells were incubated in 0.1-1 µg/mL 4,6-diamino-2-phenylindole (DAPI; ThermoFisher Scientific, D1306) for 10 minute and washed 3 times with PBS. When immunostaining was complete, the coverslip was mounted with Fluoromount-G (SouthernBiotech, 0100-01). A fluorescence microscope (Olympus, IX73P2F) was used for fluorescence imaging. The antibody used in this part was listed in Table 3.

**Table 3.** Primary and secondary antibodies.

| Antibody | Manufacturer | Cat. Number | dilution |
| --- | --- | --- | --- |
| PMEL Monoclonal Antibody (HMB45) | Invitrogen | MA513232 | 1:500 |
| Anti-TRP2/DCT | Abcam | ab74073 | 1:1000 |
| Anti-Vimentin | Abcam | ab8978 | 1:1000 |
| Melan-A Polyclonal Antibody | Invitrogen | PA5120982 | 1:1000 |
| Cytokeratin 3 Monoclonal Antibody (AE5) | Invitrogen | MA1-5763 | 1:500 |
| Cytokeratin 14 Polyclonal Antibody | Invitrogen | PA5-28002 | 1:1000 |
| Anti-p63 | Abcam | ab124762 | 1:1000 |
| Anti-Ki67 | Abcam | ab15580 | 1:1000 |
| Anti-Cytokeratin 10 | Abcam | ab9026 | 1:500 |
| Anti-P cadherin | Abcam | ab307740 | 1:500 |
| Anti-Cytokeratin 15 | Abcam | ab80522 | 1:500 |
| Human MITF Antibody | R&D systems | AF5769-SP | 1:100 |
| Human MSX2 Antibody | R&D systems | MAB7917-SP | 1:200 |
| Collagen V Polyclonal Antibody | Invitrogen | PA5-102416 | 1:250 |
| Donkey anti-Rabbit IgG, Alexa Fluor<sup>™</sup> 594 | Invitrogen | A21207 | 1:1000 |
| Donkey anti-Mouse IgG, Alexa Fluor<sup>™</sup> 488 | Invitrogen | A21202 | 1:1000 |

### Quantitative PCR

Total RNA was extracted from samples using an RNAeasy™ Animal RNA Extraction Kit (Beyotime, China) and cDNA was synthesized using a HiScript III RT SuperMix for qPCR (Vazyme, China), according to the manufacturer’s instructions. qPCR was performed on the Applied Biosystems™ 7500 RT-PCR system (ThermoFisher SCIENTIFIC, USA) and ChamQ SYBR qPCR Master Mix (Vazyme, China). The primers used in this part was listed in Table 4.

**Table 4.**
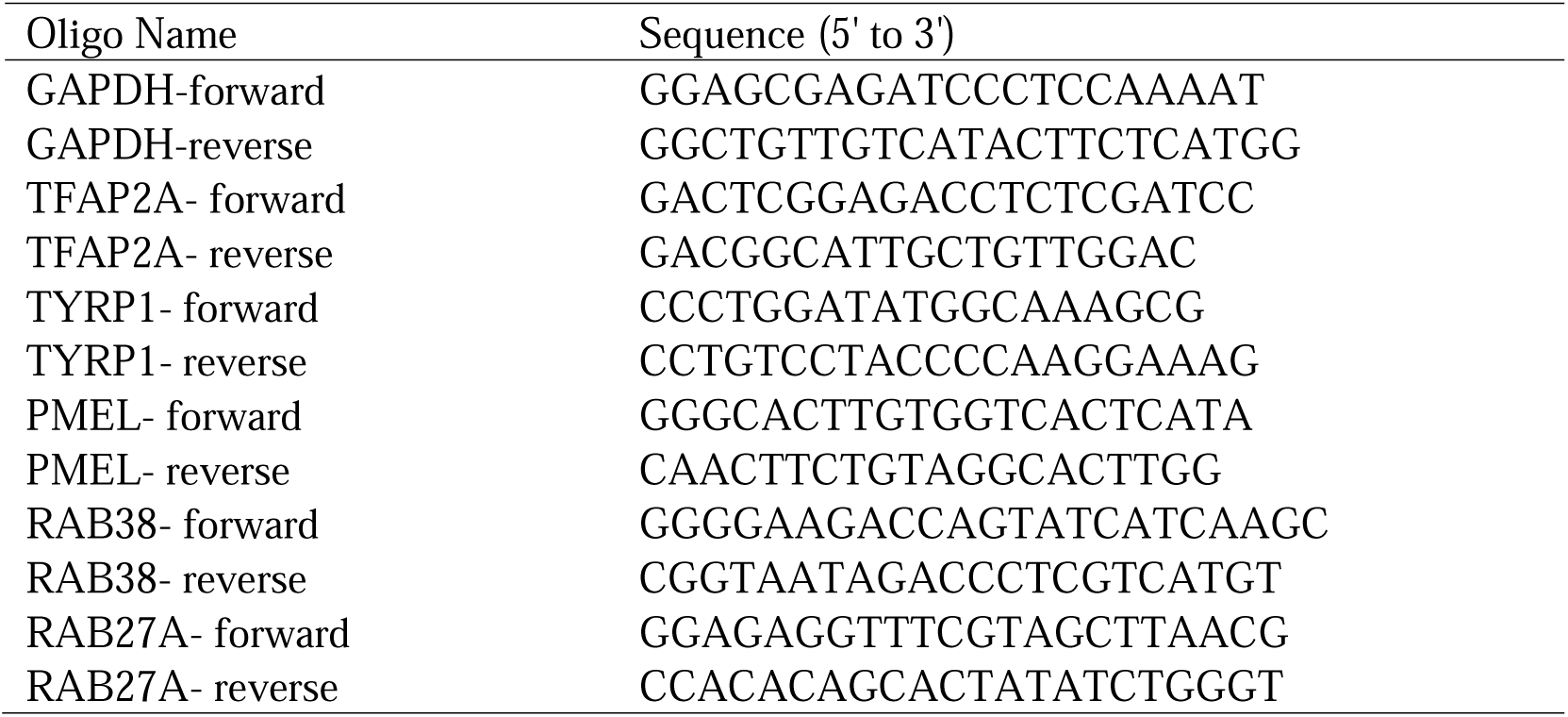
Primers for quantitative PCR.

The expression levels of all genes were normalized to that of GAPDH. Relative gene expression was determined using the 2^−ΔΔCt^ method and presented as the mean standard error of three independent experiments.

### Ethics Approval and Consent to Participate

The project named “Autologous hair follicle-derived epidermal tissue transplantation for the treatment of vitiligo” was reviewed and approved by Ethics Committee of Huashan Hospital affiliated to Fudan University as KY2020-793 on September 9, 2020. The written informed consent was obtained from all participants in this study.

### Transplantation of HFES

Transplantation of HFES were performed as described in our previous study. First, hair follicles were harvested according to lesion size and cultured separately to generate epidermal constructs. During transplantation, dermabrasion was performed on the lesions before application of cultured grafts, followed by standard dressing fixation. Dressings were removed at 2 weeks postoperatively. Patients were evaluated at 1, 3, and 6 months with documentation of repigmentation area and dermatoscopic imaging to assess recovery. Repigmentation rate (%) = (repigmented area / transplanted area) × 100. Therapeutic response rates for both techniques were statistically compared at 6 months.

### Statistical analyses

Statistical analysis of intergroup comparisons was conducted using a Student’s t-test, and results with p < 0.05 were considered statistically significant.

## Data Availability

All data produced in the present study are available upon reasonable request to the authors

## Supplementary Data

**Fig. S1.**
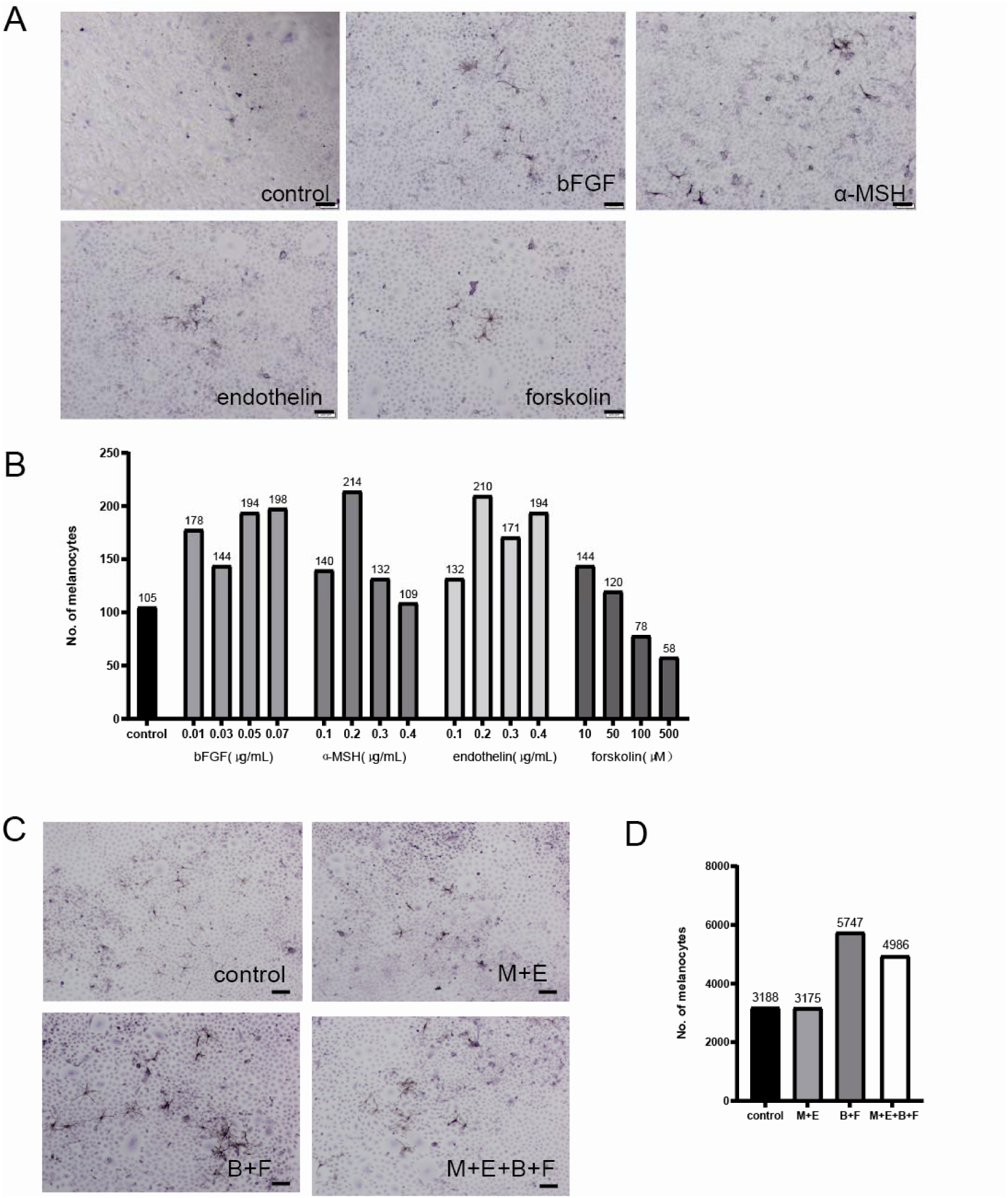
Melanocyte stimulating factors and their optimal concentrations. (A) DOPA staining image of each group with best performance. 0.05 μg/mL bFGF, 0.2 μg/mL α-MSH, 0.2 μg/mL endothelin and 10 μM forskolin. (B) Number of DOPA+ melanocytes per dish. (C) DOPA staining of combined melanocyte stimulating factors. (D) Number of DOPA+ melanocytes per dish. B:bFGF, F: forskolin, M: α-MSH, E: endothelin.

**Fig. S2.**
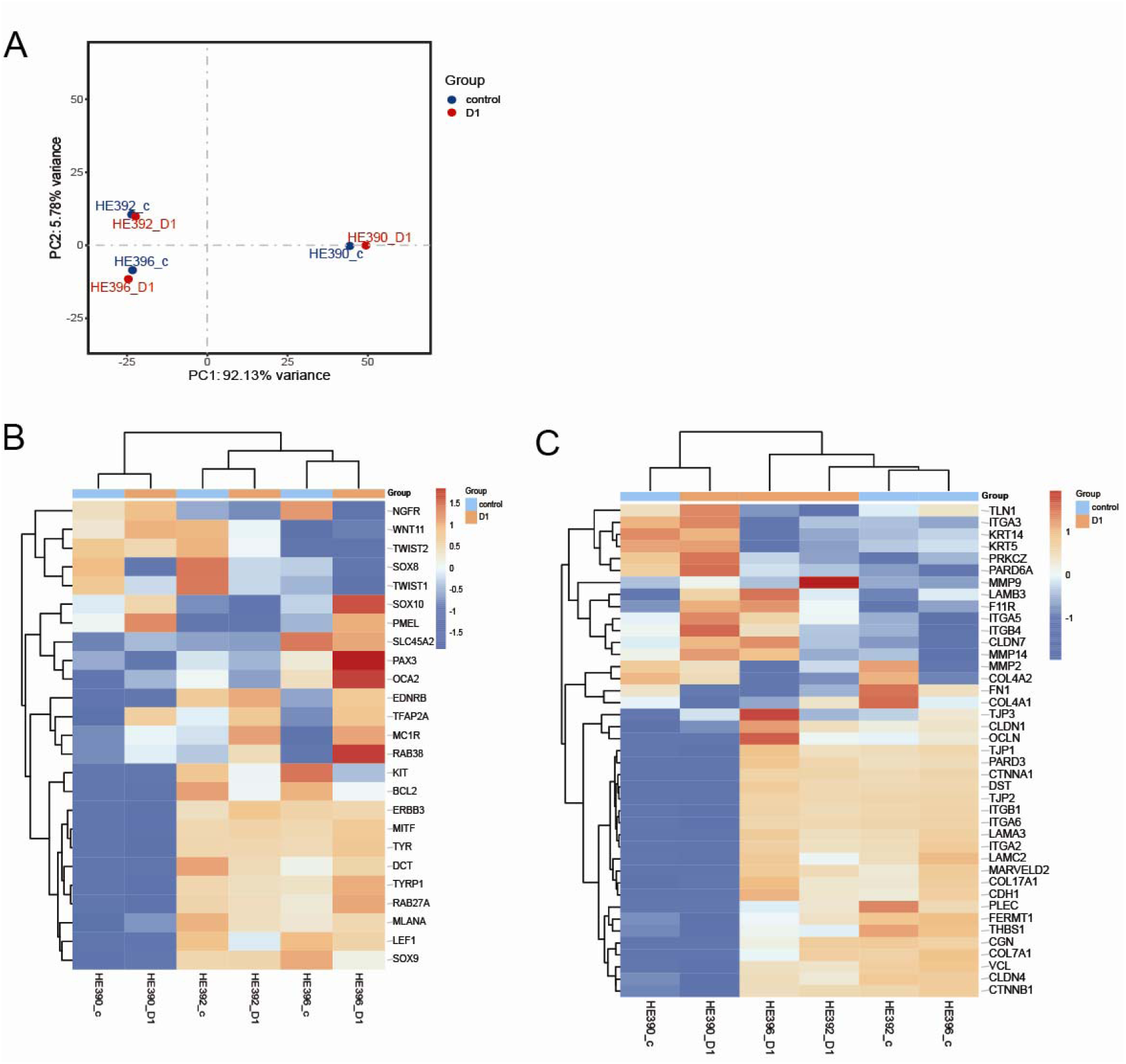
Bulk RNA sequencing comparison of cell characteristics under control and D1 culture conditions. (A) Principal-component analysis (PCA) comparing the transcriptome of control and D1. (B) Heatmap plotting expression of melanocyte related genes. (C) Heatmap plotting expression of epidermis/keratinocytes related genes.

**Table S1.**
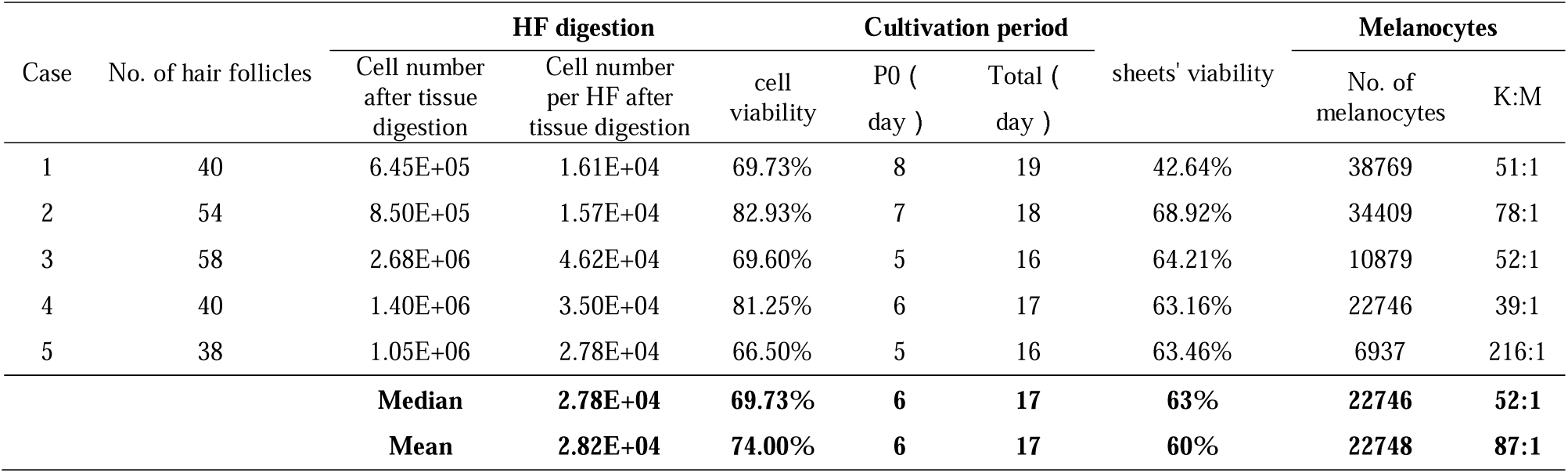
Characteristics of HFES.

**Table S2.** The clinical characteristics of the 5 enrolled patients.

| Case | Age | Gender | Race | Classification | Duration of disease | Stable period | Co-morbidities | Family history | Previous treatment |
| --- | --- | --- | --- | --- | --- | --- | --- | --- | --- |
| 1 | 38 | female | Chinese | Non-segmental | 17 years | 1 year | no | yes | Topical glucocorticoids |
| 2 | 28 | female | Chinese | Segmental | 22 years | 12 years | no | no | Topical glucocorticoids |
| 3 | 27 | male | Chinese | Segmental | 18 years | 9 years | no | no | Topical glucocorticoids |
| 4 | 32 | male | Chinese | Non-segmental | 20 years | 8 years | no | no | Topical glucocorticoids |
| 5 | 48 | male | Chinese | Non-segmental | 31 years | 6 years | no | yes | Topical glucocorticoids |

